# Modeling and Simulation of CAR T cell Therapy in Chronic Lymphocytic Leukemia Patients

**DOI:** 10.1101/2022.12.01.22282976

**Authors:** Ujwani Nukala, Marisabel Rodriguez Messan, Osman N. Yogurtcu, Zuben Sauna, Hong Yang

## Abstract

Advances in genetic engineering have made it possible to reprogram an individual’s immune cells to express receptors that recognize markers on tumor cell surfaces. The process of re-engineering T cell lymphocytes to express Chimeric Antigen Receptors (CARs) and reinfusing the CAR-modified T cells into patients to treat various cancers is being explored in clinical trials. While the majority of patients with some cancers (e.g., B cell acute lymphocytic leukemia) respond to CAR-T cell therapy, this success is not evidenced in all cancers. For example, only 26% of Chronic Lymphocytic Leukemia (CLL) patients respond to CAR T cell therapy. Understanding of the factors associated with an individual patient’s response is important for patient selection and could help develop more effective CAR T cell therapies. Here we present a mechanistic mathematical model to identify factors associated with responses to CAR T cell therapeutic interventions. The proposed model is a system of coupled ordinary differential equations designed based on known immunological principles and prevailing hypotheses on the mechanism of CAR T cell kinetics, Interleukin 6 (IL-6) secretion, and tumor killing in CAR T cell therapy. The model reports *in silico* disease outcomes using B cell concentration as a surrogate biomarker. Our results are consistent with the *in vitro* experimental observations that CAR T cell fitness in terms of its tumor cell killing capacity and proliferation plays an important role in the patient response. We demonstrate the utility of mathematical modelling in understanding the factors that play an important role in patient response to CAR T cell therapy.

## Introduction

CAR T cells are T cells that are genetically engineered to express a receptor on the T cell membrane that can recognize a specific antigen expressed on the target cancer cell, independent of the Major Histocompatibility Complex (MHC) molecules. In the presence of the target antigen, CAR T cells proliferate inside the human body and elicit an immune response against cells that express the target antigen by activating immune cells such as normal T cells and triggering the secretion of cytokines and chemokines. Currently there are six CAR T cell therapies approved by the FDA for treatment of hematological malignancies including lymphomas, leukemia, and multiple myeloma. Research on CAR T cell therapies to treat other tumors including solid tumors is in progress.

Though CAR T cell therapy has been successfully used to treat hematological malignancies, there are some uncertainties including antigen escape, wear-off of therapeutic effect and adverse events like cytokine release syndrome (CRS) and neurotoxicity. Activated CAR T cells secrete proinflammatory cytokines which facilitate tumor cell apoptosis. On the other hand, uncontrolled secretion of cytokines leads to toxicity. CRS is the most common and life-threatening adverse reaction associated with CAR T cell therapy [1]. Currently, CRS is managed using immunosuppressive drugs (e.g., tocilizumab) and/or corticosteroids [2, 3].

The treatment of Chronic Lymphocytic Leukemia (CLL) with chemotherapeutic agents such as Venetoclax and Ibrutinib results in an initial response rate of ∼95%. Unfortunately, only 10-30% of patients using these treatments achieve complete remission and 50% of the patients relapse within 3-4 years [4]. The Relapsed or Refractory (R/R) CLL patients have few treatment options.CAR T cell therapy has been demonstrated to provide a sustained response in R/R CLL patients [4, 5]. The initial response rates in CLL patients treated with CAR T cells that target the CD-19 antigen are 70-80%, but only 20-40% of the patients achieve a durable response [6]. Multiple studies have shown the promise of CAR T cell therapy for CLL patients [5, 7, 8]. However, it is not clear why some patients show a sustained remission while others do not. A better understanding of factors and variables associated with a sustained response to CAR T cell therapy would help to refine patient selection for increased efficacy.

Recently, mathematical modeling and simulation approaches have been used to study different aspects of CAR T cell therapy [2, 9-13]. The development of mathematical models describing the interplay of infused CAR T cells, cytokines, and B cells in a patient throughout the course of CAR T therapy could supplement the assessment of clinical safety and efficacy of the therapy and help with personalized dosing and more effective management of adverse events. In this study we modeled and simulated the anti-CD19+ CAR T therapy in CLL patients. The mathematical model presented here was developed to study the multiphasic kinetics of CAR T cells along with the dynamics of tumor cells and IL-6 secretion (Figure 1) and has been utilized in identifying factors that may impact the responses to CAR T cell therapy in CLL patients.

**Figure 1:**
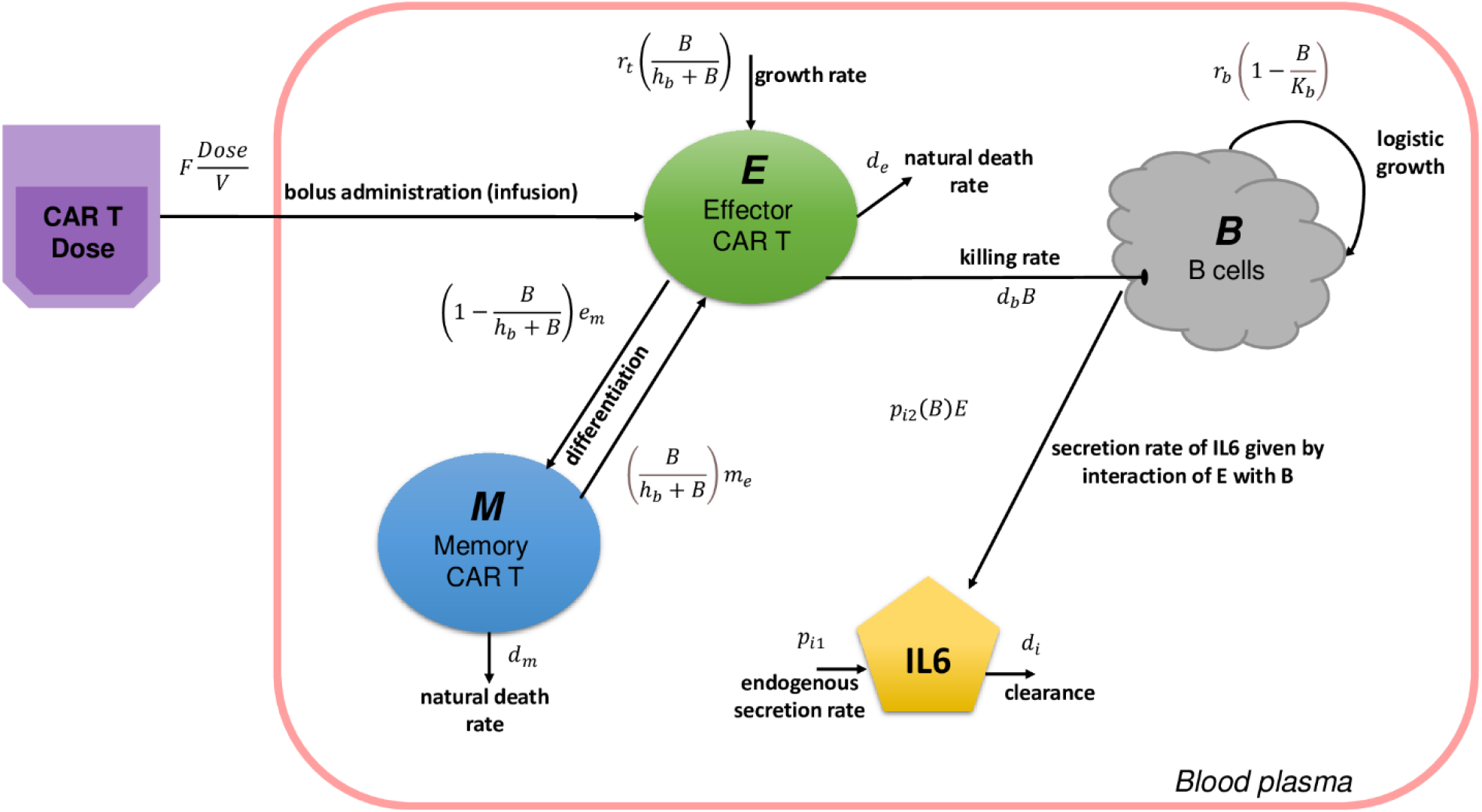
Schematic representation of the CAR T mathematical model. After infusion, only a portion of the administered dose becomes available in the blood plasma with volume ‘*V*’, and a bioavailability coefficient ‘*F*’ was considered. CAR T cells proliferate upon interaction with the tumor cells (B cells) at a maximal rate ‘*r*_*t*_’. CAR T cells interact and kill the tumor cells at a rate ‘*d*_*b*_’, upon which IL-6 is released at a rate ‘*p*_*i2*_’. Differentiation of Effector cells into Memory cells and Memory cells into Effector cells takes place based on the presence of target antigen at a rate ‘*e*_*m*_’ and ‘*m*_*e*_’ respectively. Death rates or decay constants of Effector CAR T cells, Memory CAR T cells and IL-6 are represented with ‘*d*_*e*_’, ‘*d*_*m*_’ and ‘*d*_*i*_’, respectively (Table 2).

## Methods

### Clinical data

The data used in this study, including CAR T cell concentration and IL-6 peak concentrations in the peripheral blood, collected after intravenous infusion of CTL019 in adult patients with CLL, were obtained from Porter et al. [5]. Longitudinal data reporting IL-6 levels in patients UPN1 and UPN2 were obtained from Kalos, M., et al., [9]. Longitudinal data reporting IL-6 levels in patient UPN10 were from Fraietta et al., 2018 [10]. To calibrate the model, we used patient-specific data from nine patients (including CR, PR, and NR patients, as detailed below) [5], with at least four CAR T cell observations above the quantification limit. For all patients, longitudinal data quantifying CAR T cells as copies/µg of genomic DNA obtained from supplementary Table S7 of reference [5] were converted to cells/µl following the procedure described in Mueller et al. [14].The clinical outcomes of the nine patients used in the model are categorized as Complete Remission (CR, n = 3), Partial Response (PR, n=3), and No Response (NR, n=3) based on the clinical outcomes reported in the literature [5].

### Complete Remission Patients: (Patient-UPN1, UPN2, UPN10)

Longitudinal IL-6 concentration data for Patients UPN1, UPN2 were obtained from Supplementary Table S2 and S4 (reference [9]) and data for Patient UPN10 were extracted from Fig 1b (reference [10]) using WebPlotDigitizer (https://automeris.io/WebPlotDigitizer). Baseline tumor values (cells/µl) of all the patients are estimated using the ranges extracted from Fig 1b (reference [11]) using WebPlotDigitizer, and are reported in Table 1.

**Table 1.** Patient characteristics and individual patient data used in model fitting.

| Category<br>(Based on<br>reported<br>patient<br>outcome)[5] | Patient<br>ID | Total CART<br>dose<br>infused<br>$\times 10^8$<br>cells | Initial tumor burden<br>cells/ μl<br>range extracted from<br>[11] | Baseline IL-6<br>pg/ml | IL-6 Peak<br>pg/ml |
| --- | --- | --- | --- | --- | --- |
| Complete<br>Remission<br>(CR) | UPN1 | 11.3 | 5000 - 11863 | 7[8] | 268 [4] |
|  | UPN2 | 0.142 | 5000 - 11863 | 8.7[8] | 43[4] |
|  | UPN10 | 5.61 | 5000 - 11863 | 16.2[9] | 1736[9] |
| Partial Response (PR) | UPN5 | 3.92 | 1725 - 3366 | 8.6 * | 4.8 [4] |
|  | UPN12 | 1.18 | 1725 - 3366 | 8.6 * | 369[4] |
|  | UPN22 | 0.864 | 1725 - 3366 | 8.6 * | 2607[4] |
| No Response (NR) | UPN17 | 1.03 | 10000 - 12000 | 8.6 * | 112 [4] |
|  | UPN18 | 2.77 | 10000 - 12000 | 8.6 * | 22 [4] |
|  | UPN25 | 2.71 | 10000 - 12000 | 8.6 * | 39 [4] |
| *It is the average value of reported baseline IL-6 from Patients-UPN1, UPN2 and UPN3 [8]. |  |  |  |  |  |

### Partial Response Patients (Patient-UPN5, UPN12, UPN22) and No Response Patients (Patient-UPN17, UPN18, UPN25)

Only peak IL-6 concentrations were available for these patients, which were obtained from supplementary table S11 (reference [5]). The baseline IL-6 concentration was assumed to be 8.6 pg/ml, which is the average baseline concentration reported for CLL patients in the literature [9]. The IL-6 concentration on the last day of CAR T cell observation was assumed to be the same as the baseline value (8.6 pg/ml) for all the patients, as observed in the study by Kalos et al.,2011 [9] (Table 1).

The CAR T cell data below the quantification level (<25 copies/μg DNA) were censored in the parameter estimation analysis for all the patients [5]. We assumed that all patients received three split doses (10%, 30% and 60% of the total given dose) on days 0, 1 and 2 [5], except for Patient UPN10 whose CAR T cell concentrations were reported after the second dose on day 70, and thus presumably receiving only a single dose (Table S1).

#### Model Structure

The model presented here consists of one compartment (blood plasma) and four species (i) B cells, (ii) Effector CAR T cells, (iii) Memory CAR T cells and (iv) IL-6 molecules (Figure 1). The equations used in this model were modified from Hanson [15] and Liu et al., [12]. The details of each model equation are listed below.

#### B cells (B)

B cell species include both normal and cancerous B cells and we assumed they carry the CD-19 antigen on their cell surface. In the absence of immune intervention or therapy, B cells are assumed to grow logistically, with a maximum growth rate, *r*_*b*_, and a carrying capacity, *K*_*b*_. With the intervention of immunotherapy, B cells are assumed to be killed by Effector CAR T cells at a killing rate of *d*_*b*_, which is proportional to the product of the concentration of B cells (*B*) and Effector CAR T cells (*E*). Equation 1 is used to describe the dynamics of B cells in patients treated with CD19 CAR T cell therapy.

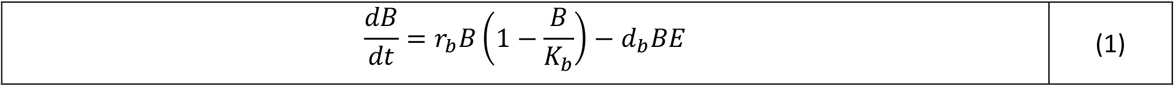

#### CAR T cells (T)

CAR T cells are administered to the patient through IV infusion, a procedure that takes approximately 5 minutes [16]. Since the time units of our model is in days, we assumed instantaneous input of CAR T cells into the blood stream at the time of administration τ in our model. Moreover, there is a transient decline of CAR T cells in the blood after the infusion, due to their distribution to other tissues. Thus, it is assumed that only a portion of the administered dose is available in the blood plasma (volume V =3 × 10^6^ µl), and that is accounted for by using a bioavailability coefficient *F* [12, 15]. Based on the observed CAR T cell concentration in the blood after each dose, *F* is estimated to range from 0.01 to 0.09 (Table S1).

After the initial infusion of CAR T cells, their concentration in the blood declines rapidly within hours because of their distribution into tissues. Following this rapid decline, the CAR T cell concentration expands to reach its maximum concentration within a few days or weeks (*T*_*max*_). This expansion phase is followed by a contraction-persistence phase where CAR T cells decline biexponentially over a variable period ranging from days to months or years [2], [12], [17],[14].

Memory cell differentiation takes place during the contraction phase resulting in the longer persistence of CAR T cells in the body. CAR T cell kinetics typically consist of four distinct phases: early distribution, expansion, contraction, and persistence [12]. However, the initial distribution phase is much shorter in CLL patients, and the contraction-persistence phases are indistinct. Consequently, CAR T cell kinetics in this study are modeled with two phases, expansion, and contraction-persistence.

The two kinetic phases of CAR T cells, the expansion phase (*t* < *T*_*max*_) and the contraction-persistence phase (*t* ≥ *T*_*max*_) are modeled [2], [12], where *T*_*max*_ is the time to reach the maximum concentration of CAR T cells. During the expansion phase (Eq. 2a), CAR T cells proliferate at a rate *rt*, triggered by the presence of B cells and the proliferation is proportional to the concentration of Effector CAR T cells. Thus, we model this mechanism with the product of a growth rate *rt*, concentration of Effector CAR T cells *E*, and a Michaelis-Menten interaction term 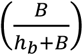, where *h*_*b*_ is the half-maximal saturation constant such that the growth of CAR T cells is 50% when *B* = *hb*. We assumed the death of Effector CAR T cells to be negligible in this phase. During the contraction-persistence phase, CAR T cells either die at a natural death rate *de* or differentiate into Memory CAR T cells (Eq. 2b; the 1^st^ and 2^nd^ term on the right). On the other hand, memory CAR T cells can also differentiate into Effector CAR T cells (Eq. 2b; the 3rd term on the right). Any of these two-way interchanges between Effector and Memory CAR T cells requires the presence of target antigen and the balance of this two-way interchange depends on the concentration of Effector and Memory CAR T cells.

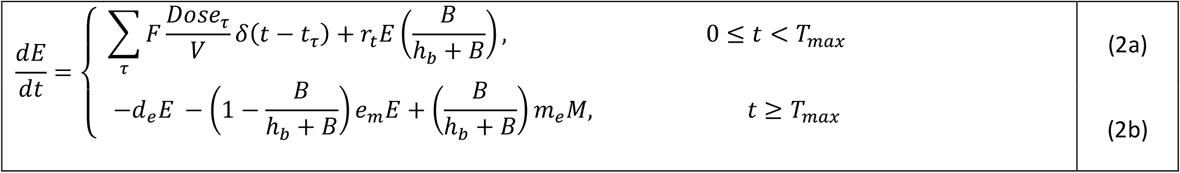

Memory CAR T cells are differentiated from Effector CAR T cells only during the contraction-persistence phase (*t ≥ T*_*max*_) and at a rate e_m_. The initial number of Memory CAR T cells at time *t* = 0 is set to M_0_ = 0, and M(t) = 0 for 0 ≤ *t* < *T*_*max*_ [12], which means that Memory CAR T cells are not differentiated from Effector CAR T cells during the expansion phase (*t* < *T*_*max*_). Like Eq 2b, the Eq3 includes two-way interchange between Effector and Memory CAR T cells (Eq3, the 1^st^ and 2^nd^ term on the right), as well as the natural death of Memory cells at a rate of *d*_*m*_(the 3^rd^ term on the right).

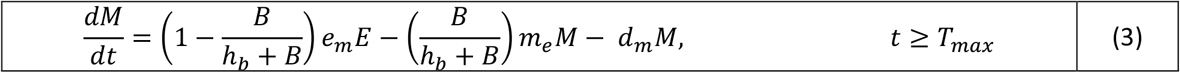

#### IL-6 molecules (IL-6)

IL-6 molecules are assumed to have a baseline endogenous secretion rate *p*_*i*_1 and are eliminated at a rate *d*_*i*_. The baseline values of IL-6 are patient specific and *p*_*i*_1 is estimated as the product of IL-6_0_ and *d*_*i*_, where IL-6_0_ is the initial concentration of IL-6 molecules. IL-6 is secreted at a rate *p*_*i*_2 by CAR T cells when they interact with B cells, as described in Equation 4.

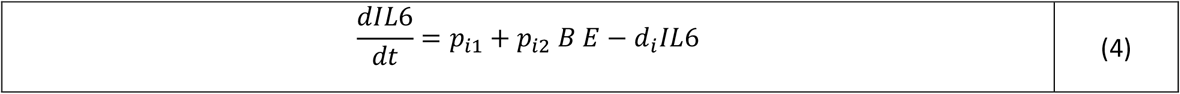

### Model parameters and data fitting

The model consists of 14 parameters (Table 2), of which nine are estimated by fitting the model to observed individual patient data and the other five parameter values were derived from the literature. Since patients are categorized into CR, PR, and NR groups, three sets of parameters were estimated along with inter-individual variability. The range of initial tumor cell concentration B_0_ was extracted from Figure 1b (reference [11]), as shown in Table 1, and the initial tumor cell concentration for each patient response group was estimated via model fitting.

**Table 2.**
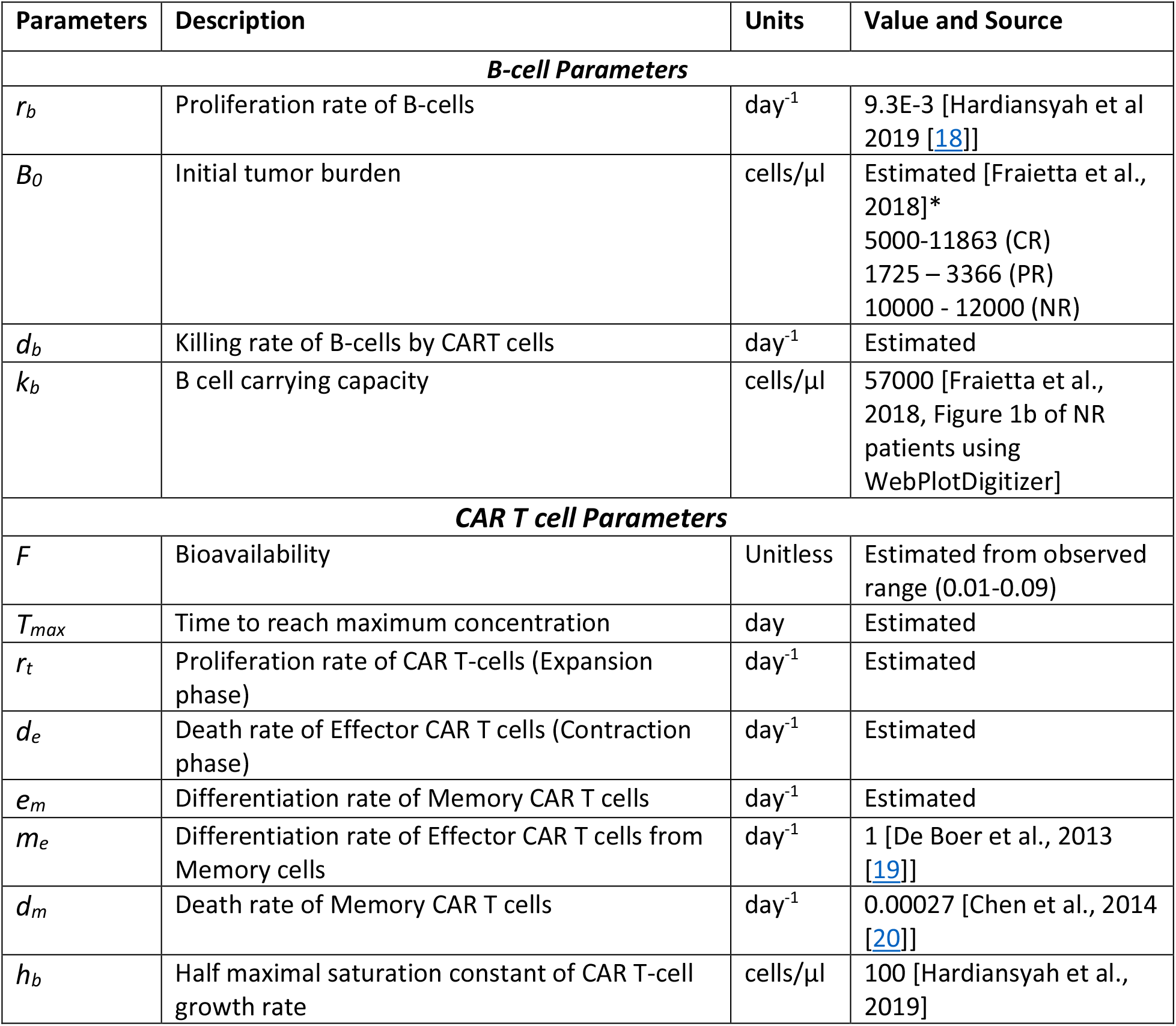
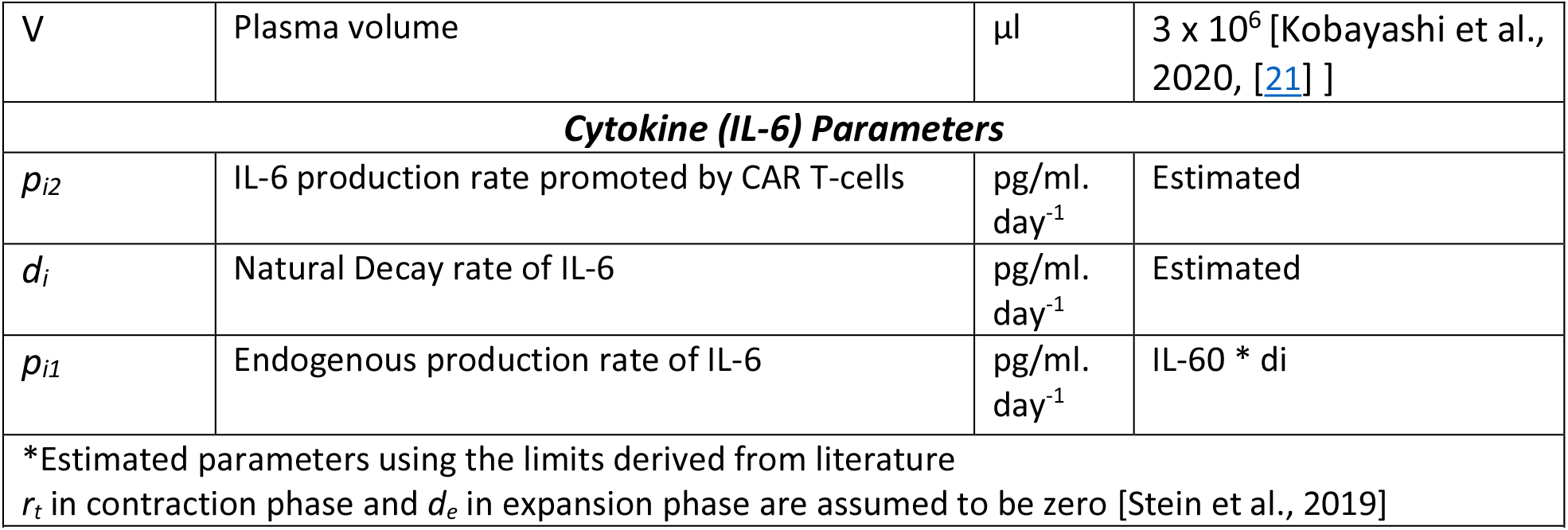
Model Parameters

| Parameters | Description | Units | Value and Source |
| --- | --- | --- | --- |
| ***B-cell Parameters*** |  |  |  |
| $r_b$ | Proliferation rate of B-cells | day<sup>-1</sup> | 9.3E-3 [Hardiansyah et al 2019 [18]] |
| $B_0$ | Initial tumor burden | cells/μl | Estimated [Fraietta et al., 2018]*<br>5000-11863 (CR)<br>1725 – 3366 (PR)<br>10000 - 12000 (NR) |
| $d_b$ | Killing rate of B-cells by CART cells | day<sup>-1</sup> | Estimated |
| $k_b$ | B cell carrying capacity | cells/μl | 57000 [Fraietta et al., 2018, Figure 1b of NR patients using WebPlotDigitizer] |
| ***CAR T cell Parameters*** |  |  |  |
| $F$ | Bioavailability | Unitless | Estimated from observed range (0.01-0.09) |
| $T_{max}$ | Time to reach maximum concentration | day | Estimated |
| $r_t$ | Proliferation rate of CAR T-cells (Expansion phase) | day<sup>-1</sup> | Estimated |
| $d_e$ | Death rate of Effector CAR T cells (Contraction phase) | day<sup>-1</sup> | Estimated |
| $e_m$ | Differentiation rate of Memory CAR T cells | day<sup>-1</sup> | Estimated |
| $m_e$ | Differentiation rate of Effector CAR T cells from Memory cells | day<sup>-1</sup> | 1 [De Boer et al., 2013 [19]] |
| $d_m$ | Death rate of Memory CAR T cells | day<sup>-1</sup> | 0.00027 [Chen et al., 2014 [20]] |
| $h_b$ | Half maximal saturation constant of CAR T-cell growth rate | cells/μl | 100 [Hardiansyah et al., 2019] |
| V | Plasma volume | $\mu\text{l}$ | $3 \times 10^6$ [Kobayashi et al., 2020, [21] ] |
| **Cytokine (IL-6) Parameters** |  |  |  |
| $p_{i2}$ | IL-6 production rate promoted by CAR T-cells | pg/ml.<br>day<sup>-1</sup> | Estimated |
| $d_i$ | Natural Decay rate of IL-6 | pg/ml.<br>day<sup>-1</sup> | Estimated |
| $p_{i1}$ | Endogenous production rate of IL-6 | pg/ml.<br>day<sup>-1</sup> | IL-60 * $d_i$ |
| *Estimated parameters using the limits derived from literature<br>$r_t$ in contraction phase and $d_e$ in expansion phase are assumed to be zero [Stein et al., 2019] | | | |
| Table 2. Model Parameters |  |  |  |

Population pharmacokinetic analyses were performed in Monolix version 2020R1 ((**lixoft.com**) using the stochastic approximation expectation (SAEM) algorithm for nonlinear mixed-effect modeling. The parameters estimated from fitting the data are listed in Table 2. Random effects were also modeled on these parameters assuming log-normal distributed variance, and a proportional residual error model was used for CAR T cells, and IL-6, with normal distribution of residuals. The model was evaluated based on the goodness of fit plots comparing observed versus model predicted CAR T cell concentrations (Supplementary Figure 1) and reasonable Relative Standard Error (RSE%) and inter-individual variability (IIV) values.

### Sensitivity Analysis

To identify the parameters differentiating responders (CR) from non-responders (NR), parameter sensitivity analysis was carried out on five parameters (including d_b_, d_e_, e_m_, r_t_, T_max_) by testing one parameter at a time. These five parameters were selected, on the assumption that they determine the fitness of CAR T cells and thus they may be responsible for patient response. In this analysis, 1,000 virtual patients each were generated for the CR and NR groups, via random sampling using the estimated population parameters and their inter-individual variability. Using Simulx 2020R1, Monte Carlo simulations were performed in virtual CR patients, with each of the five parameters replaced one by one, with the corresponding parameter estimated for NR group, and the proportion of virtual patients achieving CR and NR response after the parameter replacement, was determined. A similar set of simulations were performed on 1,000 NR virtual patients by replacing one parameter at a time with CR parameter estimates. The sample size (N=1,000) was chosen where stabilized median model prediction for CAR T and IL-6 concentrations were achieved. Graphs were plotted, and statistical tests were performed using R version 4.1.0.

## Results

### Fitting

The model accurately simulated the kinetics of CAR T cells and IL-6 in nine CLL patients who were grouped into CR, PR, or NR response groups. We fitted 57 CAR T cell and 32 IL-6 observations in CR patients, 34 CAR T cell and 9 IL-6 observations in NR patients, and 43 CAR T cell and 9 IL-6 observations in PR patients (Figures 2-4).

**Figure 2.**
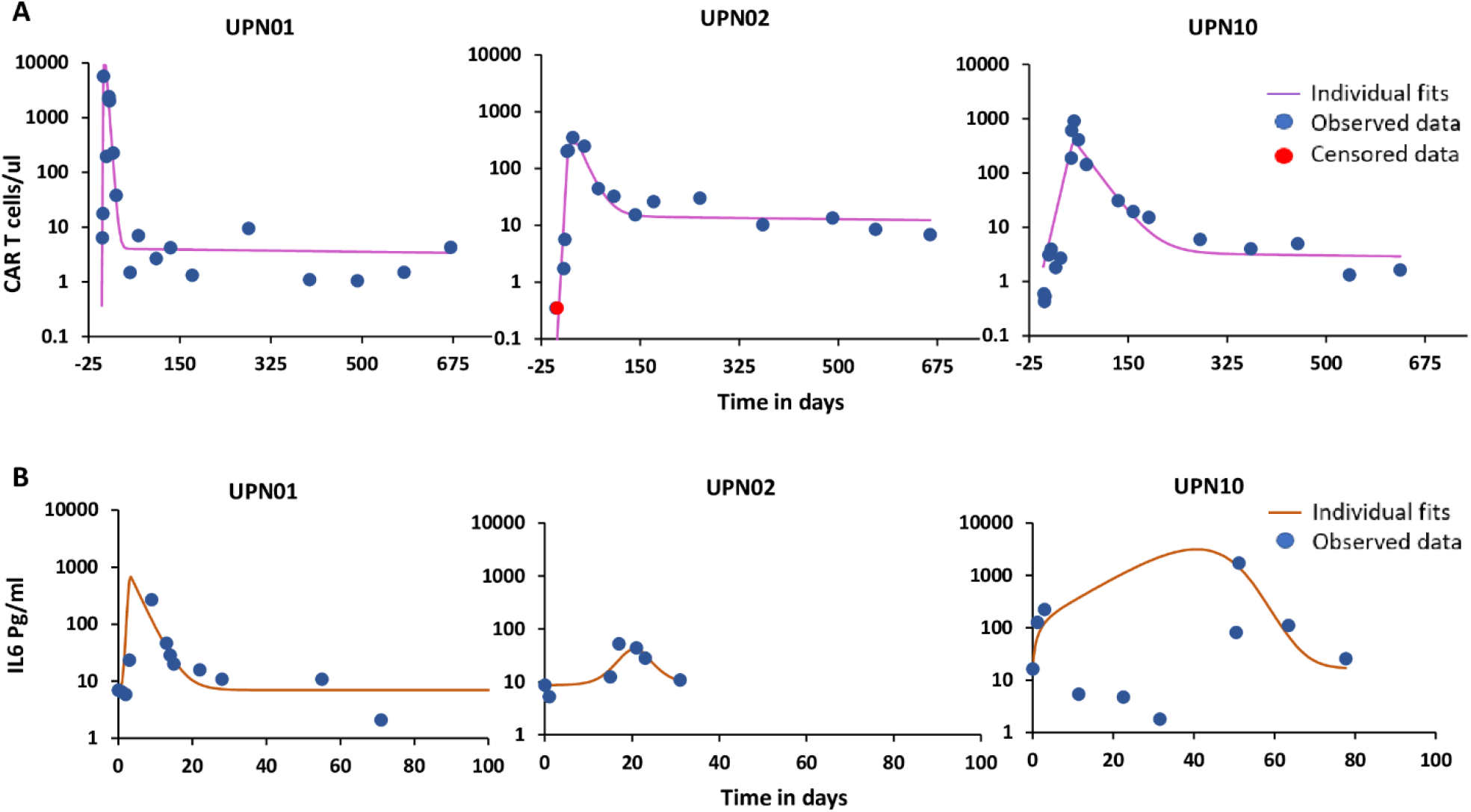
Model fittings of patients reported as Complete Remission. (A) Individual patient data fitting of CAR T cells (B) Individual patient data fitting of IL-6. Blue dots are the observations, red dots are observations below the detection limit for CAR T cells and the solid line is the model prediction. Note: Data for IL-6 is available for different time points in each patient and the data points do not correspond to CAR T cell data points.

**Figure 3.**
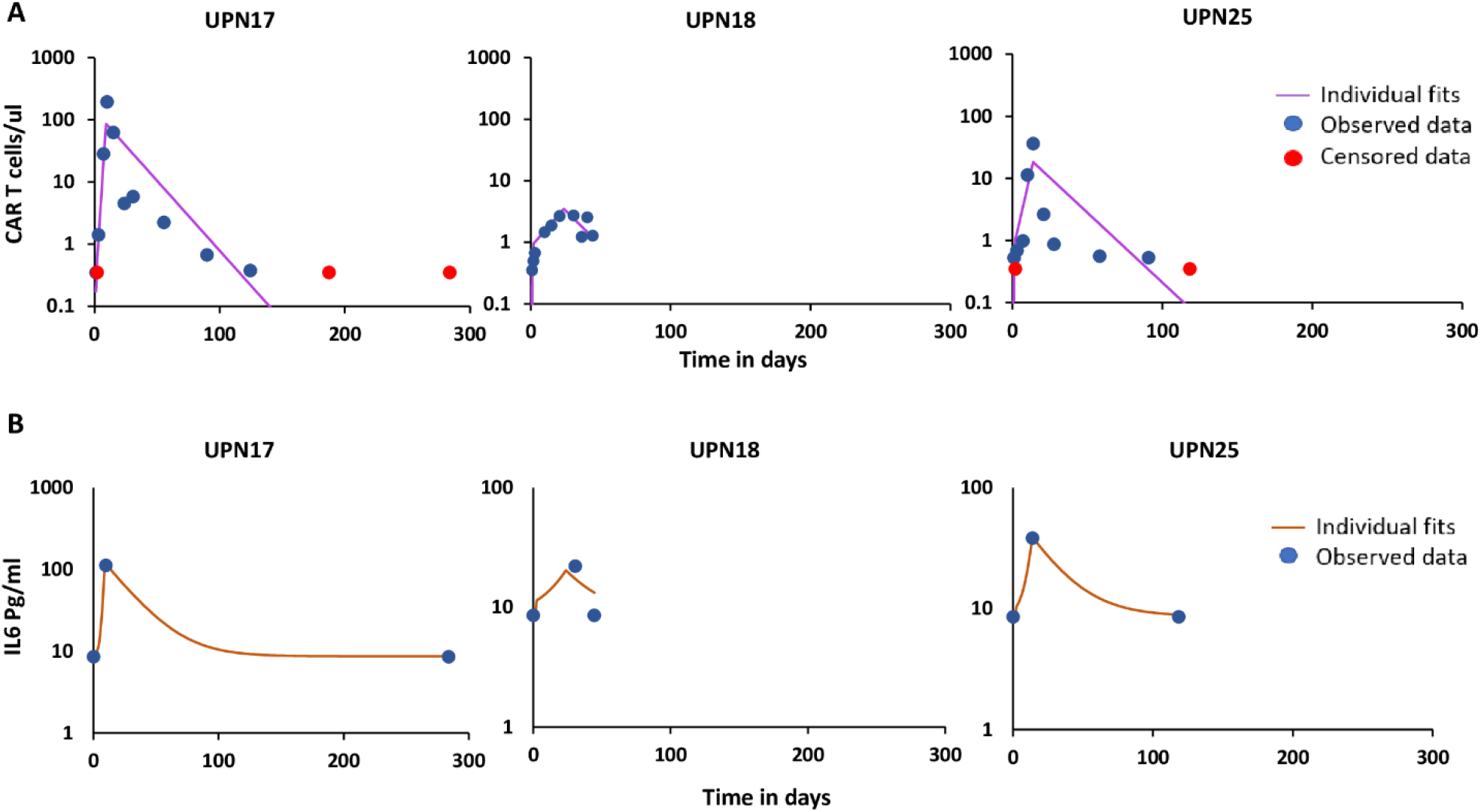
Model fittings of patients reported as No Response. (A) Individual patient data fitting of CAR T cells (B) Individual patient data fitting of IL-6. Blue dots are the observations, red dots are observations below the detection limit for CAR T cells and the solid line is the model prediction. Note: Data for IL-6 is available for different time points in each patient and the data points do not correspond to CAR T cell data points.

**Figure 4.**
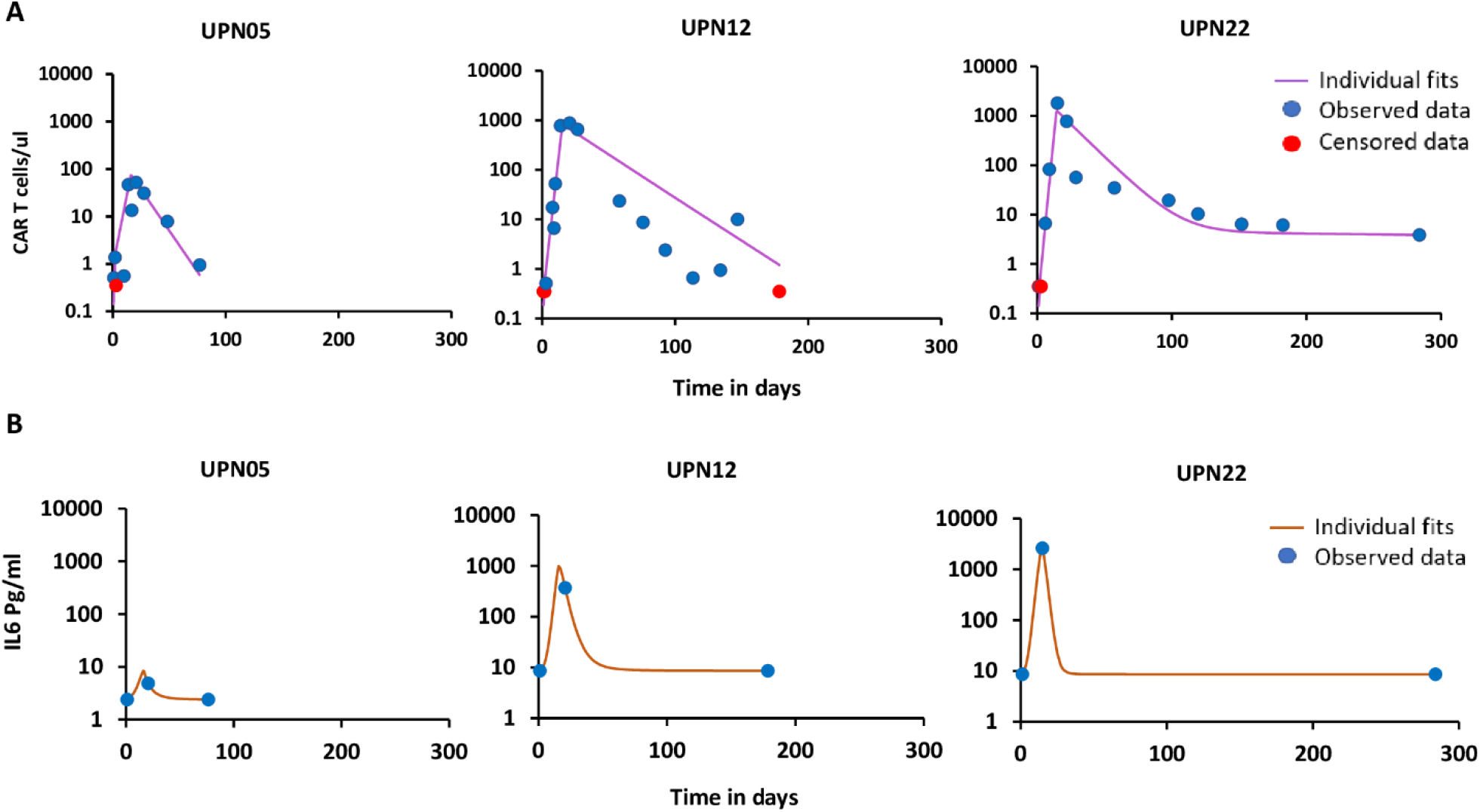
Model fittings of patients reported as Partial Response. (A) Individual patient data fitting of CAR T cells (B) Individual patient data fitting of IL-6. Blue dots are the observations, red dots are observations below the detection limit for CAR T cells and the solid line is the model prediction. Note: Data for IL-6 is available for different time points in each patient and the data points do not correspond to CAR T cell data points.

The population parameters estimated showed reasonable variability and %RSE (Table 3). Nevertheless, since the sample size is small (three patients per response group) and the observed data sparse, %RSE and IIV are large for some of the parameters. However, the model performed well, which is evidenced by the predictions of B cell concentrations in each group (Figure 5). In this study, we used B cell concentration as a surrogate biomarker of patient response, as CD-19 surface antigen, expressed on B cells is the CAR T cell target, used in this clinical trial and B cell aplasia was seen in patients achieving CR[5] (Figure 5). The model’s predictions of B cell concentrations align well with the reported patient response in each response group. In CR patients, B cells reach concentrations that are below the limit of detection in all three patients. In NR patients, B cell concentrations increase until they reach carrying capacity. Finally, in PR patients, B cell concentrations increase after a transient decline. Moreover, compared to CR patients, NR patients showed low levels of Memory CAR T cell differentiation (Figure 6) which could be a contributing factor to the long-term remission in CR patients [11]. Though PR patients showed Memory CAR T cell differentiation (Figure 6), relapse in these patients may be attributed to tumor silencing or modification of target antigen which is often seen [22]; However, these attributes are not included in the current model and the PR group is not included in the virtual population simulations, which are discussed in the sensitivity analysis section.

**Table 3.**
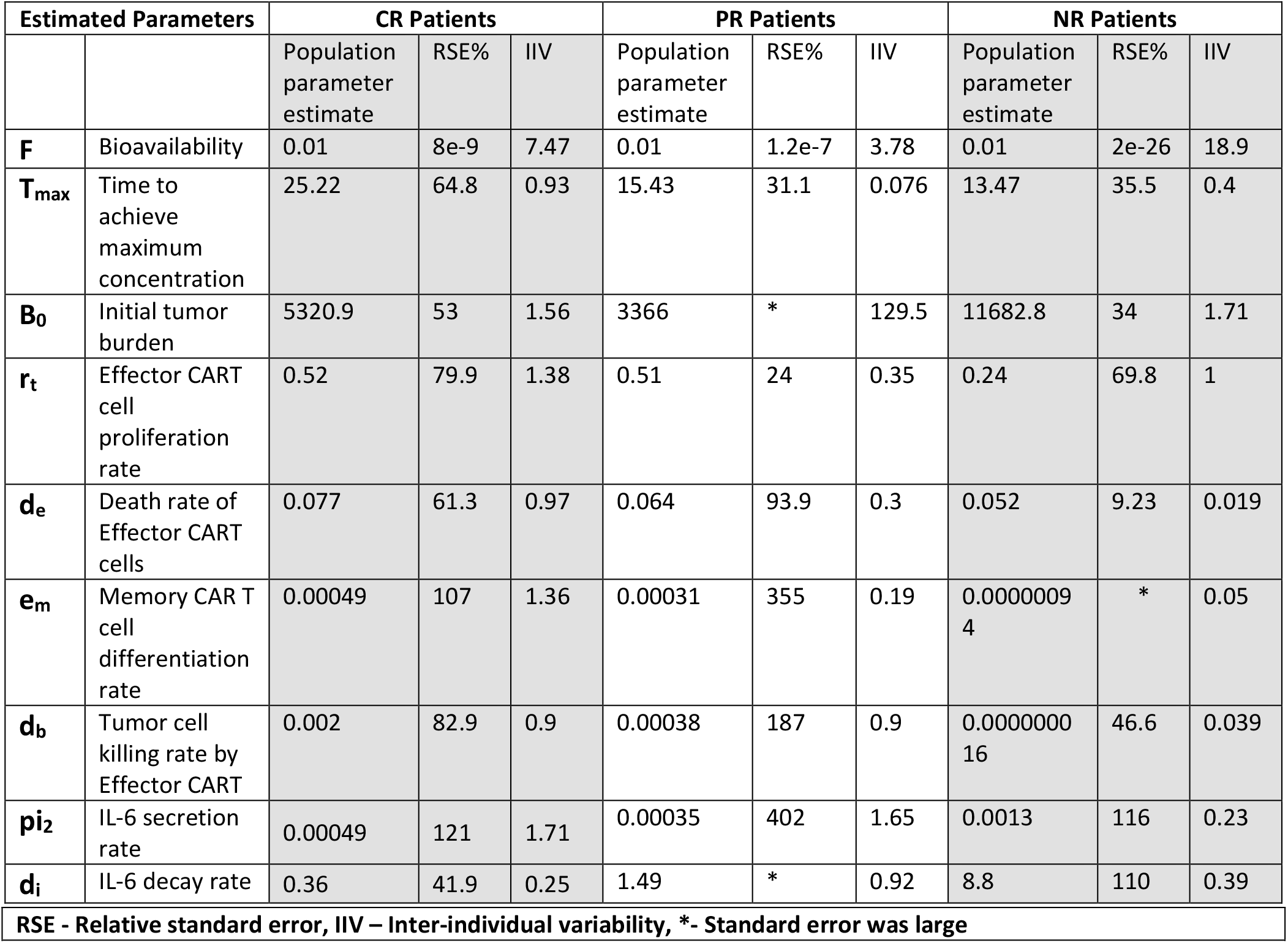
Estimated population parameters in CR, PR, and NR patients

| Estimated Parameters |  | CR Patients |  |  | PR Patients |  |  | NR Patients |  |  |
| --- | --- | --- | --- | --- | --- | --- | --- | --- | --- | --- |
|  |  | Population parameter estimate | RSE% | IIV | Population parameter estimate | RSE% | IIV | Population parameter estimate | RSE% | IIV |
| **F** | Bioavailability | 0.01 | 8e-9 | 7.47 | 0.01 | 1.2e-7 | 3.78 | 0.01 | 2e-26 | 18.9 |
| **T<sub>max</sub>** | Time to achieve maximum concentration | 25.22 | 64.8 | 0.93 | 15.43 | 31.1 | 0.076 | 13.47 | 35.5 | 0.4 |
| **B<sub>0</sub>** | Initial tumor burden | 5320.9 | 53 | 1.56 | 3366 | * | 129.5 | 11682.8 | 34 | 1.71 |
| **r<sub>t</sub>** | Effector CART cell proliferation rate | 0.52 | 79.9 | 1.38 | 0.51 | 24 | 0.35 | 0.24 | 69.8 | 1 |
| **d<sub>e</sub>** | Death rate of Effector CART cells | 0.077 | 61.3 | 0.97 | 0.064 | 93.9 | 0.3 | 0.052 | 9.23 | 0.019 |
| **e<sub>m</sub>** | Memory CAR T cell differentiation rate | 0.00049 | 107 | 1.36 | 0.00031 | 355 | 0.19 | 0.00000094 | * | 0.05 |
| **d<sub>b</sub>** | Tumor cell killing rate by Effector CART | 0.002 | 82.9 | 0.9 | 0.00038 | 187 | 0.9 | 0.000000016 | 46.6 | 0.039 |
| **pi<sub>2</sub>** | IL-6 secretion rate | 0.00049 | 121 | 1.71 | 0.00035 | 402 | 1.65 | 0.0013 | 116 | 0.23 |
| **d<sub>i</sub>** | IL-6 decay rate | 0.36 | 41.9 | 0.25 | 1.49 | * | 0.92 | 8.8 | 110 | 0.39 |

|  |
| --- |
| RSE - Relative standard error, IIV – Inter-individual variability, *- Standard error was large |
| Table 3. Estimated population parameters in CR, PR, and NR patients |

**Figure 5.**
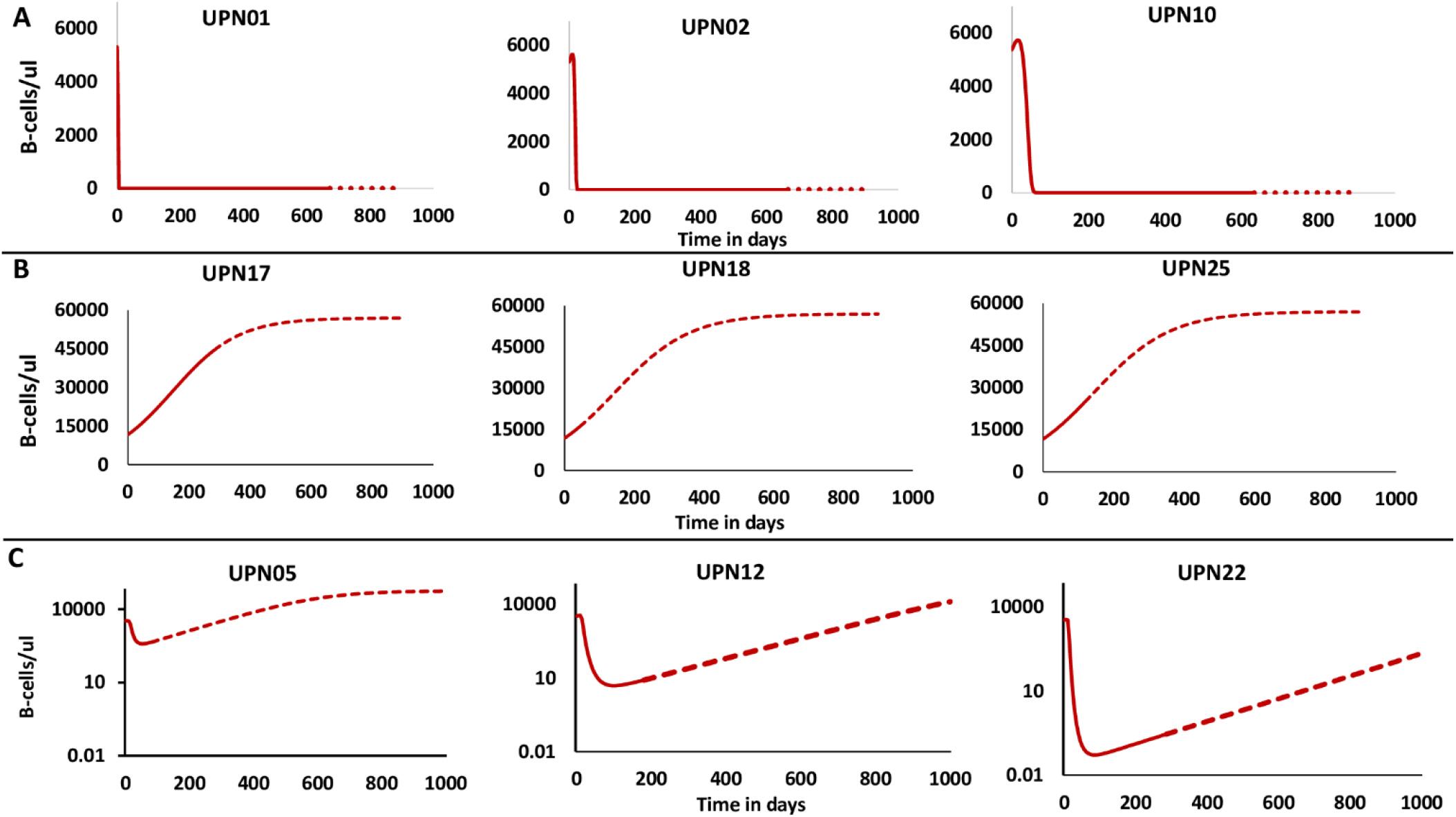
Tumor cell model predictions in (A) CR patients, (B) NR patients, and (C) PR patients. The solid line corresponds to the period of clinical study and the dotted line corresponds to the post-clinical study period.

**Figure 6.**
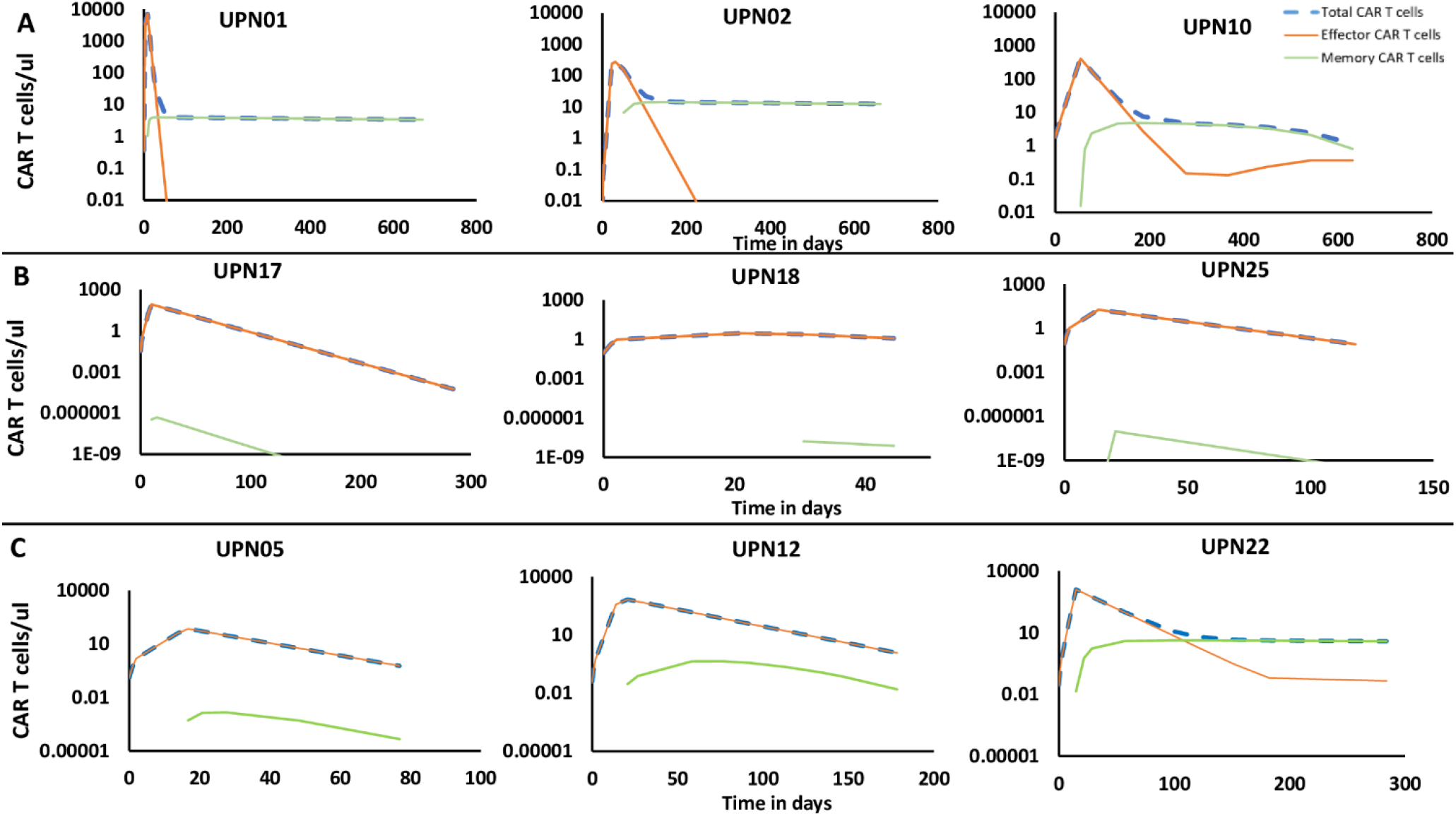
Model predictions of CAR T cell sub types in (A) CR Patients (B) NR Patients and (C) PR Patients for the duration of clinical trial period.

### Model Validation

The ability of the final calibrated model to predict the kinetics of CAR T cells, IL-6 and tumor cells was evaluated in patients UPN09, UPN06, UPN07 and UPN14 for whose CAR T cell dosage, CAR T cell numbers and IL-6 peak concentrations have been reported [5]. Since patient UPN09 achieved complete remission, population parameters estimated from the CR group with IIV and a 3-day split dosage of reported CAR T cells were used to predict the dynamics of CAR T cells, IL-6 and B cells. Patients UPN06, UPN07 and UPN14 were reported to be NR, thus population parameters estimated from the NR group with IIV were used to predict dynamics of CAR T cells, IL-6 and B cells. The observed data fell within the prediction interval for CAR T cells, IL-6, and B cells in UPN09 patient but because of the sparse observed data and most of the observed data points are below the detection levels in patients UPN06, UPN07 and UPN14, the prediction intervals are wide and not all the observed data points fell within the prediction interval (Supplementary Figure 2). This is one of the limitations of the model, and as more individual patient longitudinal data become available, the performance of the model may be improved. The validated model was then used to perform a sensitivity analysis to identify the factors that are associated with patient response.

### Sensitivity Analysis

Response to CAR T cell therapy may differ based on characteristics related to the patient, disease, or treatment regimen. In our model, some of the patient specific characteristics are captured by specific model parameters for each response group. For example, in this study, response to CAR T cell therapy was measured in terms of B cell concentrations as a surrogate biomarker for response. A virtual population of 1,000 patients in each CR and NR response groups were generated using their respective estimated population parameters and their interindividual variability. These virtual patients may characterize the heterogeneity in real world patient cohorts and sensitivity analysis may help identify the parameters that are most influential on patient response. Violin plots in Figure 7 shows the distribution and the density of each population parameter between CR and NR virtual patients; all of which are significantly different between the response groups (Wilcoxon test, p<0.05).

**Figure 7.**
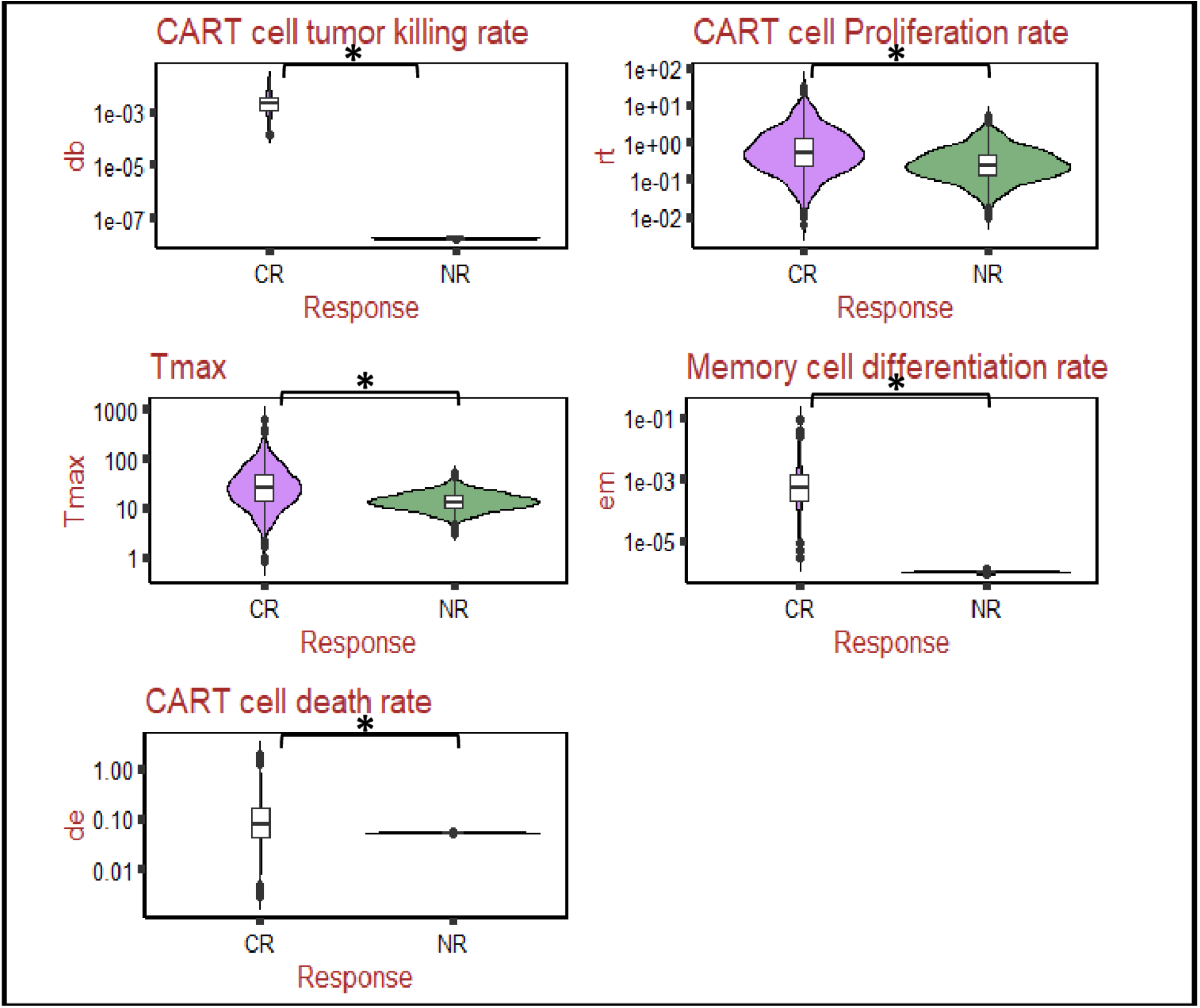
Violin plot shows the comparison between Complete Remission patient and No Response patient parameter distributions and density. All the parameters are found to be statistically significantly different between the two groups (Wilcoxon test, p<0.05).

Each parameter in CR virtual patients is replaced by the corresponding mean parameter value estimated for the NR response group and vice-versa. The CAR T cell therapy response is evaluated in the virtual patients over a 60-day period. In our simulations, the patient’s response is classified as CR when the number of tumor cells is less than or equal to one B cell/µl. This assumption is consistent with-a previous report [23], where a patient was classified as CR in the absence of Minimal Residual Disease. Minimal Residual Disease was defined as the concentration of CLL in the blood or marrow of <1 cell per 10,000 leukocytes and the normal leukocyte range being 4500-11000/µl. We assume one B cell/µl as a threshold to classify the patients as CR. A patient is classified as NR when the number of B cells is >1 B cell/µl. As shown in Figure 8A, varying the parameter *d*_*b*_ representing the tumor cell killing rate by CAR T cells, in CR virtual patients results in more NR patients compared to the other parameters. Whereas *r*_*t*_ and *T*_*max*_ showed moderate influence on patient response and the least impact was seen when *e*_*m*_ or *d*_*e*_ is replaced. Similar results were seen in the NR group when each parameter is replaced with the CR mean parameter. Figure 8B shows the percent change in the patient response (CR to NR or NR to CR) on day 60 for the virtual populations of CR and NR patients. This percent change is greatest when the parameter *d*_*b*_ is replaced, followed by *r*_*t*_, and *T*_*max*_. Replacement of the parameters *e*_*m*_ and d_e_ had the least effect on the percent change from CR to NR (or vice versa). The median tumor cell dynamics of the virtual patients when one parameter is replaced at a time, over a 180-day period is shown in Supplementary Figure 3. Though the parameter *e*_*m*_, representing the rate at which Effector CAR T cell differentiates into Memory CAR T cells, does not show an impact on the patient response, the persistent remission seen in CR patients may be attributed to the presence of Memory CAR T cells. Figure 7 shows that *e*_*m*_ is greater in CR patients compared to NR patients.

**Figure 8.**
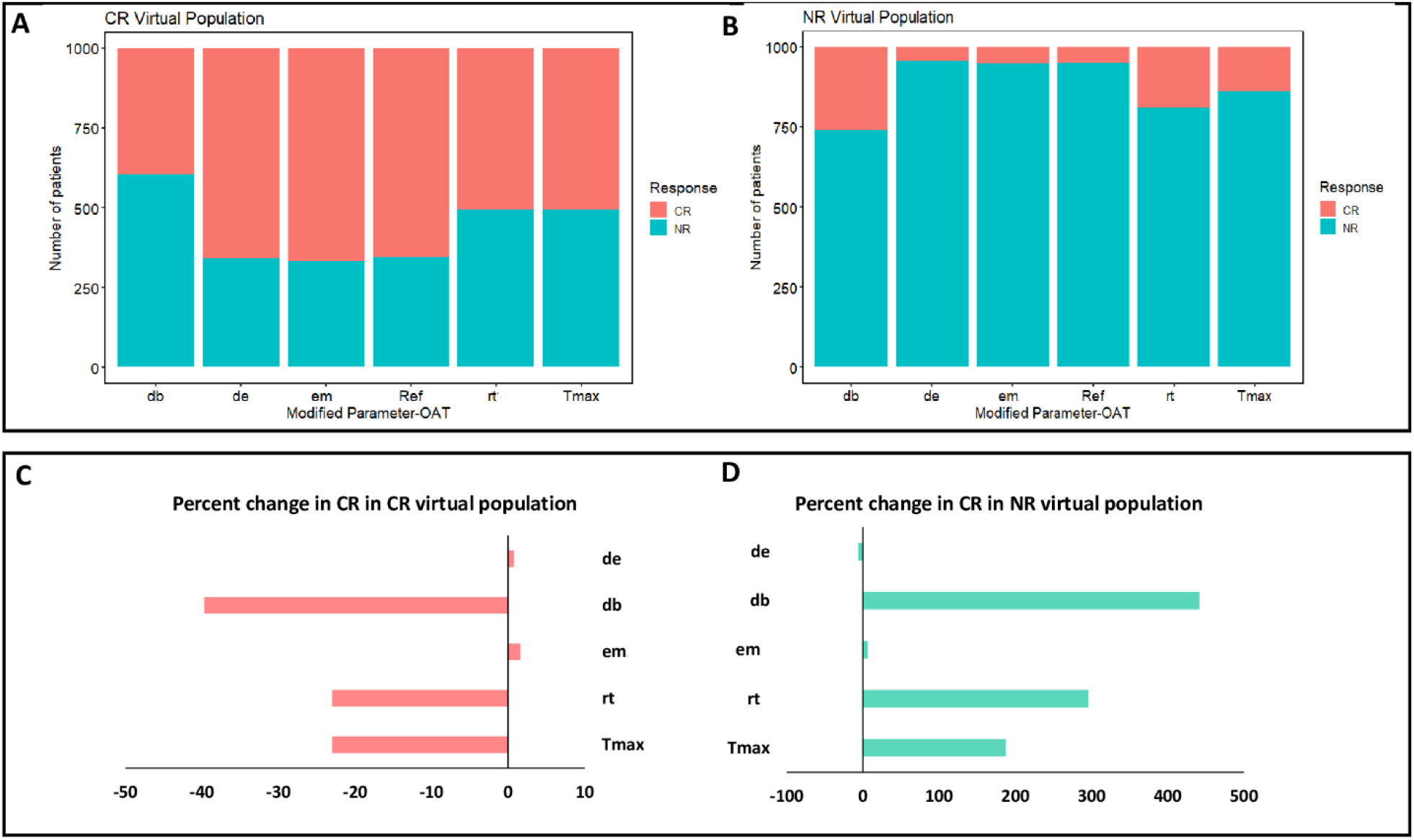
Virtual population simulations showing the impact on patient response when parameters are changed one at a time (OAT). (A) Number of patients classified as Complete Remission or No Response when one parameter is varied at a time in Complete Remission virtual patients. (B) Number of patients classified as Complete Remission or No Response when one parameter is varied at a time in No Response virtual patients. (C) Percent change in patients achieving Complete Remission among Complete Remission virtual patients when each of the parameter is varied. (D) Percent change in patients achieving Complete Remission among No Response virtual patients when each of the parameter is varied.

### Parameter correlations

Supplementary Figure 4 shows the results of a correlation analysis among the model parameters. CAR T cell proliferation rate (*r*_*t*_) is slightly negatively correlated with *T*_*max*_ and initial tumor burden (B_0_). *T*_*max*_ is slightly positively correlated with initial tumor burden, and Memory CAR T cell differentiation rate (*e*_*m*_) is slightly negatively correlated with CAR T cell proliferation rate (*r*_*t*_) and tumor killing rate (*d*_*b*_). However, none of these correlations were significant except a small negative correlation between CAR T cell proliferation rate (*r*_*t*_) and initial tumor burden (*B*_*0*_) in NR group.

## Discussion

CLL is the most common type of leukemia in adults. Despite significant advances in treatment options in recent years, the majority of patients are still incurable [24]. CAR T cell therapy is efficacious in treating many patients with leukemias and lymphomas. The USFDA has approved six CAR T cell therapies to treat various hematological malignancies. Moreover, CAR T cell therapies have the potential to treat or manage other cancers, including solid tumors [25].Multiple clinical trials conducted to study the effectiveness of CAR T cells in treating CLL have demonstrated that the therapy results in a complete response in some patients [5], [7], [26],[27].However, a significant proportion of the patients relapse.

The response rates for patients with CLL and Acute Lymphoblastic Leukemia (ALL) when treated with CAR T cells are also very different. Although the identical manufacturing process is used to manufacture CAR T cells for use in treating CLL and ALL patients, the patients with CLL show a lower remission rate. The lower response rate of CLL patients has been attributed to the functional defects and diminished proliferation capacity of T cells [22],[24], as well as the relatively older age of CLL patients [5], [22], [26], immune dysregulation because of the chronic disease evidenced by hypogammaglobulinemia, increased susceptibility to infections and autoimmune anemias [4], [24], and local immunological inhibition [28].

Previous studies were not able to identify patient specific factors such as age, prior therapies, peripheral tumor burden, p53 status, or disease specific factors that could accurately predict the response of an individual CLL patient to CAR T cell therapy [5],[11]. However, patients who responded to the therapy exhibited dramatic in vivo CAR T cell expansion in the first two weeks after infusion followed by log-normal decay in peripheral blood, and CAR T cells persisted over a long term, which is coincident with B cell aplasia. On the other hand, non-responders exhibited limited or no in vivo CAR T cell expansion and a limited degree of B cell aplasia in the first six months after infusion [11]. These results underscore the importance of CAR T cell functionality in terms of their in vivo expansion, and persistence with ongoing anti-tumor activity for long-term remissions.

Our results, using a mathematical model, are consistent with previous clinical studies which show that the response to CAR T cell therapy in CLL patients is influenced by intrinsic T cell fitness. The mechanistic basis of why some patients have T cells that are optimally functional while others do not, requires further study. Prior therapies could provide one explanation. For instance, a recent study showed that previous treatment with ibrutinib helps CAR T cells efficiently kill cancer cells in CLL patients [22]. Studies carried out to elucidate the factors responsible for patient response to the CAR T cell therapy are beset with several challenges, which are detailed in our previous study [29]. Studies performed in one population, may not be applicable to other patient populations because of the heterogeneity in the patient specific factors, disease characteristics, and the CAR T product characteristics. The dose given to each patient is different. Additionally, the study population sizes are considerably small, and the inter-individual variability is large. Though several studies attempt to identify the covariates associated with patient response post-infusion, it is of immense importance to identify a biomarker associated with patient response as well as adverse events such as CRS, before the infusion of CAR T cells. In this regard, several studies have proposed that ex vivo CAR T cell proliferation potential is associated with in vivo proliferation upon antigen stimulation [22]. CAR T cell characteristics including the pre-infusion *in vitro* proliferation and tumor cell killing rates could be potential biomarkers of patient response [11],[30]. Clinical data show that CRS occurs before CAR T cells reach their peak concentration [26]. However, CAR T cell expansion capacity is associated with CRS, hence pre-infusion CAR T cell characteristics may be an optimal predictor of CRS. However, this postulate also needs further investigation. Collectively, the current state of the art suggests that the identification of pre-infusion biomarkers is a clear unmet need for the selection of patients who would benefit from CAR T cell therapy. Such a biomarker could also be useful in designing clinical strategies for those patients who are currently non-responsive to CAR T cell-based therapies.

In this study, our aim was to use mathematical modeling as a tool to study the cellular kinetics of CAR T cells and identify the factors that drive patient response. Specifically, this model framework helps to understand how the features related to CAR T cell fitness, including the tumor cell killing rate (*d*_*b*_), proliferation rate (*r*_*t*_), and proliferation duration (*T*_*max*_) influence the patient response to CAR T cell therapy. Since the data on CAR T cell subsets (CD4^+^/CD8^+^) and phenotypes (Naïve, Central memory, Effector memory cells) in the infused product were not publicly available, it was assumed that the infused CAR T cells consisted of only Effector CAR T cells. We modeled Effector CAR T cells such that they differentiate to Memory CAR T cells after reaching a maximum concentration at *T*_*max*_ to distinguish expansion and contraction-persistent phases, though Memory CAR T cell differentiation may occur in the expansion phase as well. The Memory CAR T cells formed may differentiate back to Effector CAR T cells upon target engagement in the persistence phase. Although in addition to IL-6, IFNγ, TNF, IL-2, C-reactive protein, and ferritin were reported as potential biomarkers of CRS [31], the majority of the publications only report IL-6 levels. This is based on evidence that IL-6 is a key driver of CRS [32]. Our model also includes only IL-6. However, were more data to be available, our model could easily be amended to include additional cytokines.

Our model was calibrated using experimentally determined data on CAR T cell and IL-6 concentrations. Individual patient data were fitted to the model based on the reported response to estimate population parameters for each response group, including CR, PR, and NR. The multiphasic CAR T cell kinetics were recapitulated by our model (Figures 2-4), and the therapeutic responses reported as predictions of B cell concentration (Figure 5) align with the clinical responses of individual patients. The model presented here captured clinical outcomes (CR, PR, and NR) depending on group-specific model parameters. This demonsstrates that the model can capture responses that depend on patient variability. Patient response was closely associated with CAR T cell kinetics. Patients who achieved CR had high CAR T proliferation rates (*r*_*t*_), *T*_*max*_, and lower contraction rates (*d*_*e*_) compared to NR patients. The Memory CAR T cell differentiation rate and tumor cell killing rate are also significantly higher in CR patients than in NR patients (Figure 7).

Parameter sensitivity analysis was performed to identify the parameters that distinguish responders from non-responders, by testing patient-specific measurables such as tumor cell killing rate of CAR T cells (*d*_*b*_), the rate of CAR T cell proliferation (*r*_*t*_), time to reach maximum CAR T cell concentration (*T*_*max*_), CAR T cell contraction rate (*d*_*e*_) and Memory CAR T cell differentiation rate (*e*_*m*_). Our sensitivity analysis showed that the tumor cell killing rate of CAR T cells (*d*_*b*_) has the largest impact on the patient response. Replacing *d*_*b*_ parameter in NR group with CR parameter group resulted in greater increase in CR response, indicating that change in effectiveness of CAR T cell anti-tumor activity may affect therapeutic outcome. The next most influential parameters are *r*_*t*_, the CAR T cell proliferation rate, and *T*_*max*_, time to reach maximum concentration, which also relates to CAR T cell fitness. These results are consistent with those reported in other studies [11],[22], in which the intrinsic fitness of CAR T cells is a significant factor in disease responsiveness. Though there may be other factors which may drive the therapy outcome, such as pre-conditioning, dose, and tumor burden, CAR T cell fitness has been shown to be an important factor for complete remission [11].

There are multiple limitations to our model, mainly arising from the limited sample size and sparse observed data, including high RSE% and IIV for some of the estimated parameters. However, it was observed that CLL patients show a high CV% for *T*_*max*_, AUC_(0-28)_ [14] and the estimated IIV in another study was high in CLL patients compared to other types of cancers [12].Though some of the patients are given tocilizumab, it is assumed in our model that it does not affect CAR T cell kinetics [2]. Tumor cell concentration in the blood is used as a biomarker for the response. This is only a surrogate biomarker of the total tumor burden.

The strengths of this model include systematically capturing patient heterogeneity, identifying parameters differentiating different response groups, and making predictions using a virtual population that may provide a better understanding of the uncertainties and heterogeneity in real-world patients.

In summary, employing CAR T cell therapy to modulate the immune responses in CLL patients is a key step, where the immunodeficiency associated with CLL is worsened by the current therapies. To optimize CAR T cell therapies in CLL, there remain many variables that require additional study. These include, the best time to infuse CART cells, pre-conditioning regimen [22],[33], CAR T cell costimulatory domain [34], target antigen, and combination of CAR T and immune checkpoint inhibitors [5],[24]. With the availability of CAR T cell phenotypic data and longitudinal cytokine data, the model we present here can be extended to study the kinetics of CAR T cell phenotypes, cytokines, and their effect on therapeutic outcome. These findings could determine important CAR T cell product characteristics before the infusion of CAR T cells to achieve improved efficacy. Our model may also be used to identify whether a candidate patient is likely to respond to CAR T cell therapy. Applying this therapy only to those patients most likely to respond would allow patients who are unlikely to respond to be treated with other available therapies.

## Data Availability

All data produced in the present work and the model code are contained in the manuscript and supplementary material

## Acknowledgments

We thank, Yin Huang Ph.D., Hussein Ezzeldin Ph.D., and Barbee Whitaker, Ph.D. for reviewing the manuscript.

## Supporting Documents

**Table S1**. Experimental data of CART and IL-6 used in model fitting

**Supplementary document S2**. Additional supporting figures and Model code

## Notes

**CONFLICT OF INTEREST** The authors declare there is no conflict of interest. This article reflects the views of the authors and should not be construed to represent the FDA’s views or policies. These comments do not bind or obligate FDA.

**FUNDING** This project was supported in part by an appointment to the Research Participation Program at OBPV/CBER, U.S. Food and Drug Administration, administered by the Oak Ridge Institute for Science and Education through an interagency agreement between the U.S. Department of Energy and the FDA.

### Competing Interest Statement

The authors have declared no competing interest.

### Funding Statement

This project was supported in part by an appointment to the Research Participation Program at OBPV/CBER, U.S. Food and Drug Administration, administered by the Oak Ridge Institute for Science and Education through an interagency agreement between the U.S. Department of Energy and the FDA.

### Author Declarations

Data used in this study is publicly available before the initiation of this study. Below are the links for all the datasets used in this study. Porter et al., 2015 - Table S7 and S11 - https://www.science.org/doi/10.1126/scitranslmed.aac5415#supplementary-materials Kalos et al., 2011 - Table S2 and S4 - https://www.science.org/doi/10.1126/scitranslmed.3002842#supplementary-materials Fraietta et al., 2018 - Figure 1b is digitized using WebPlotDigitizer - https://www.ncbi.nlm.nih.gov/pmc/articles/PMC6320248/#!po=2.50000

