## Supplementary Document-2 for "Modeling and Simulation of CAR T cell Therapy in Chronic Lymphocytic Leukemia Patients"

### Supplementary Figures:

**A**

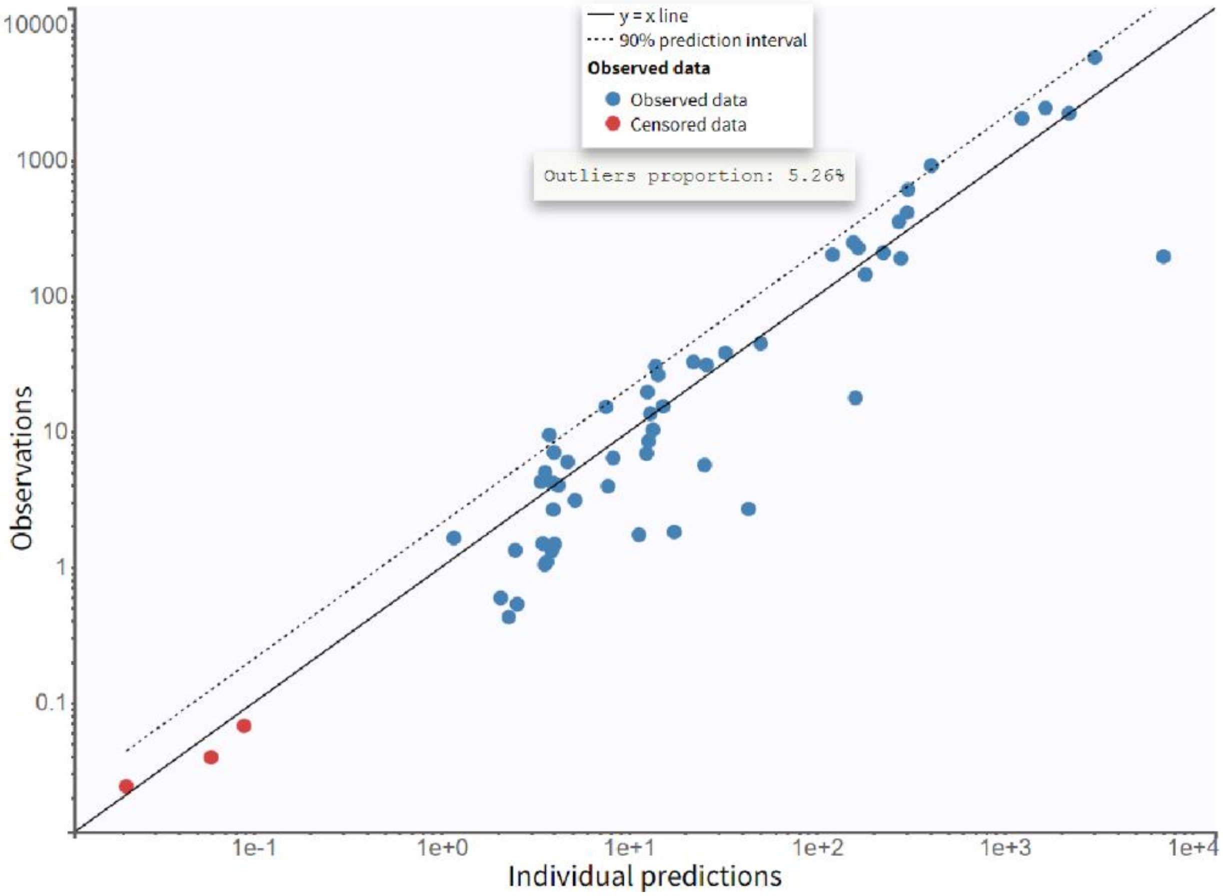

B

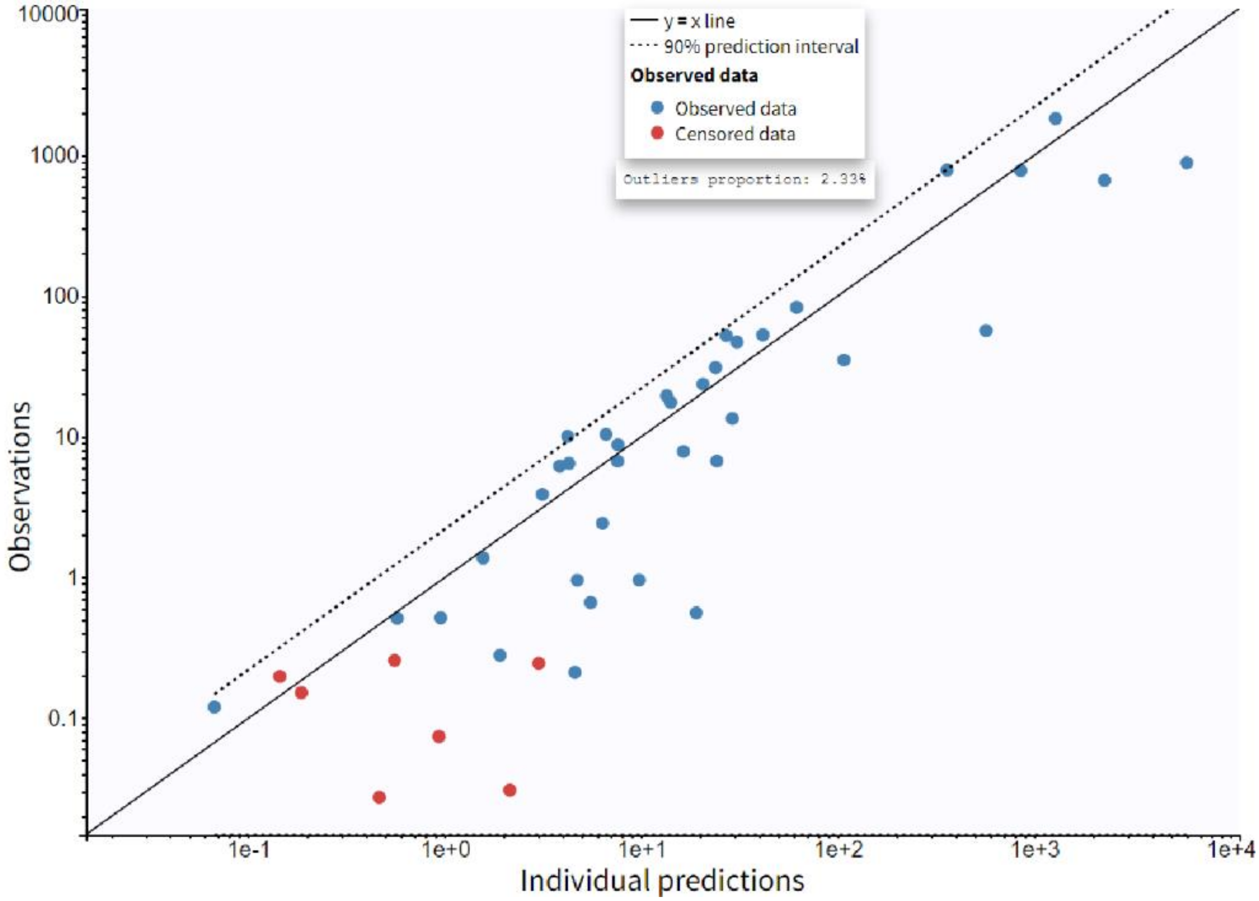

y = x line
90% prediction interval
Observed data
Observed data
Censored data
Outliers proportion: 2.33%
10000
1000
100
10
1
0.1
Observations
1e-1
1e+0
1e+1
1e+2
1e+3
1e+4
Individual predictions

C

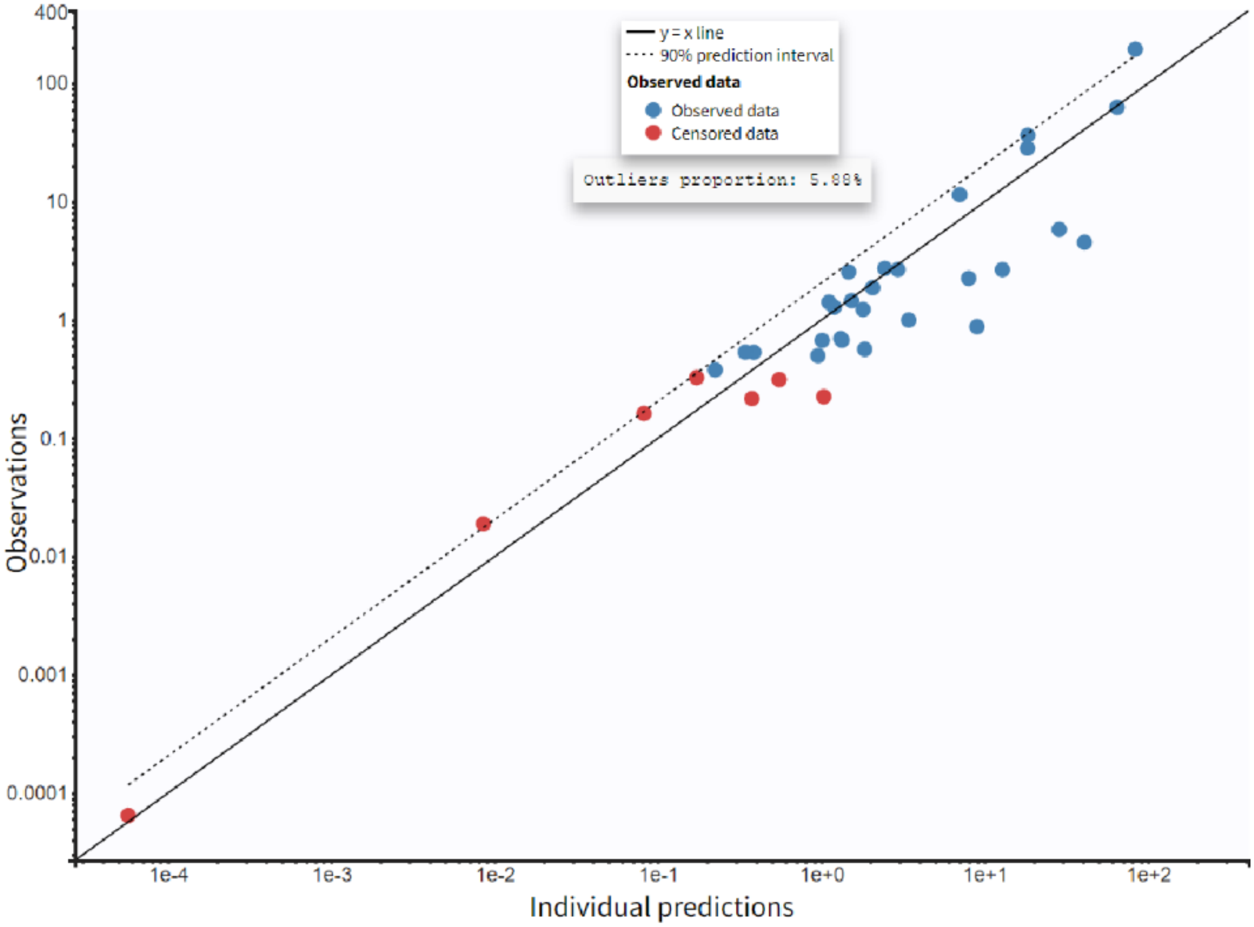


**Supplementary figure 1.** Goodness-of-fit plots showing observed versus model predicted individual data points of CAR T cells in (A) CR patients (B) PR patients and (C) NR patients

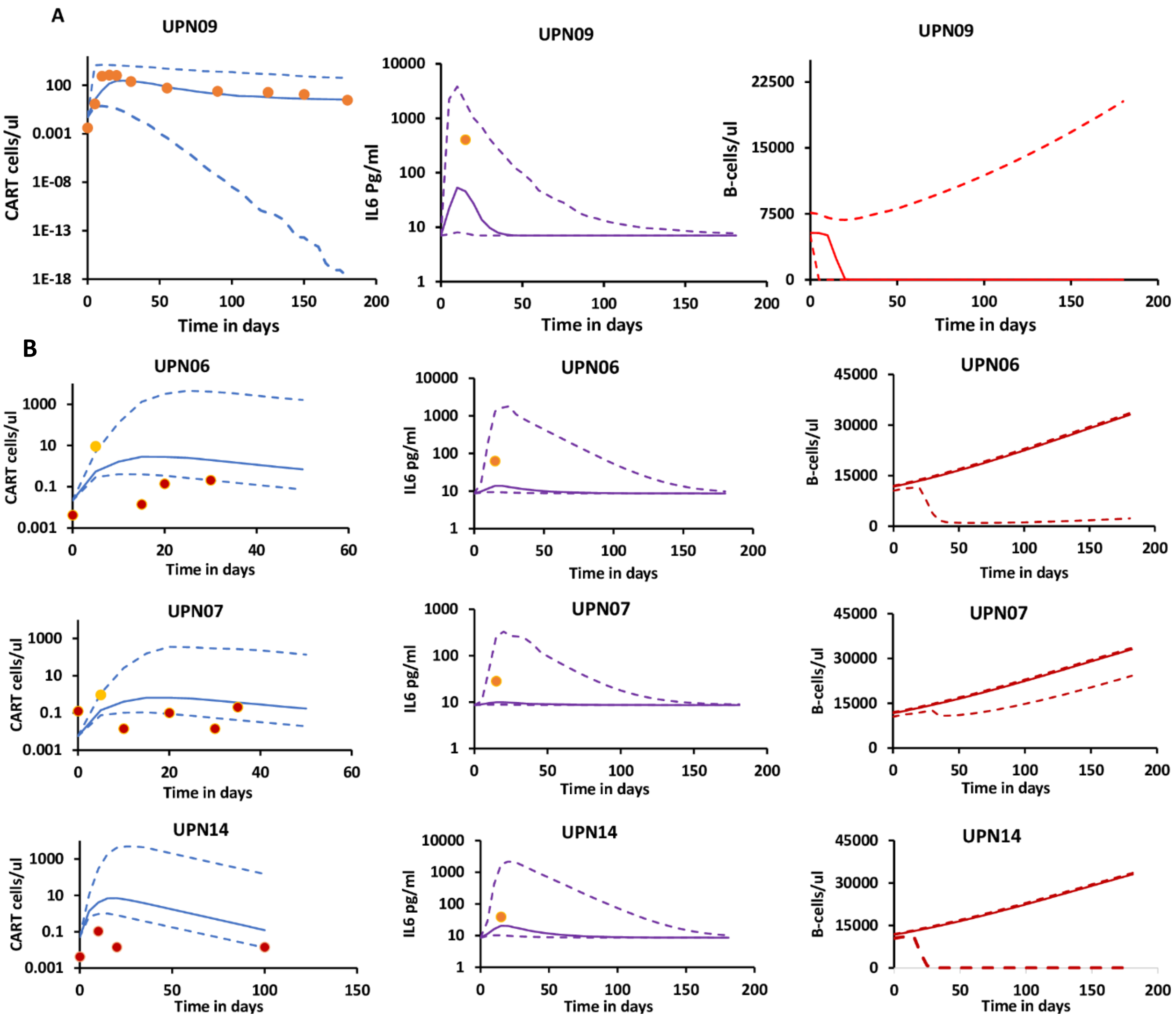


**Supplementary figure 2.** (A) Model validation plots showing model predictions for CAR T cells, IL6 and Tumor cells in patient (UPN09) reported as CR. Solid dots are observed data and Solid line is model predicted median with 95% CI as dotted lines. Observed data for B cells was not available and B cell concentration predictions are used as surrogate for patient response. (B) Model validation plots showing model predictions for CAR T cells, IL6 and Tumor cells in patients (UPN06, UPN07, UPN14) reported as NR. Solid dots are observed data and Solid line is model predicted median with 75% CI for CART, IL6 and 95% CI for B-cells as dotted lines. For patients UPN06, UPN07, UPN14, red color solid dots for CAR T cells are below the detection level.

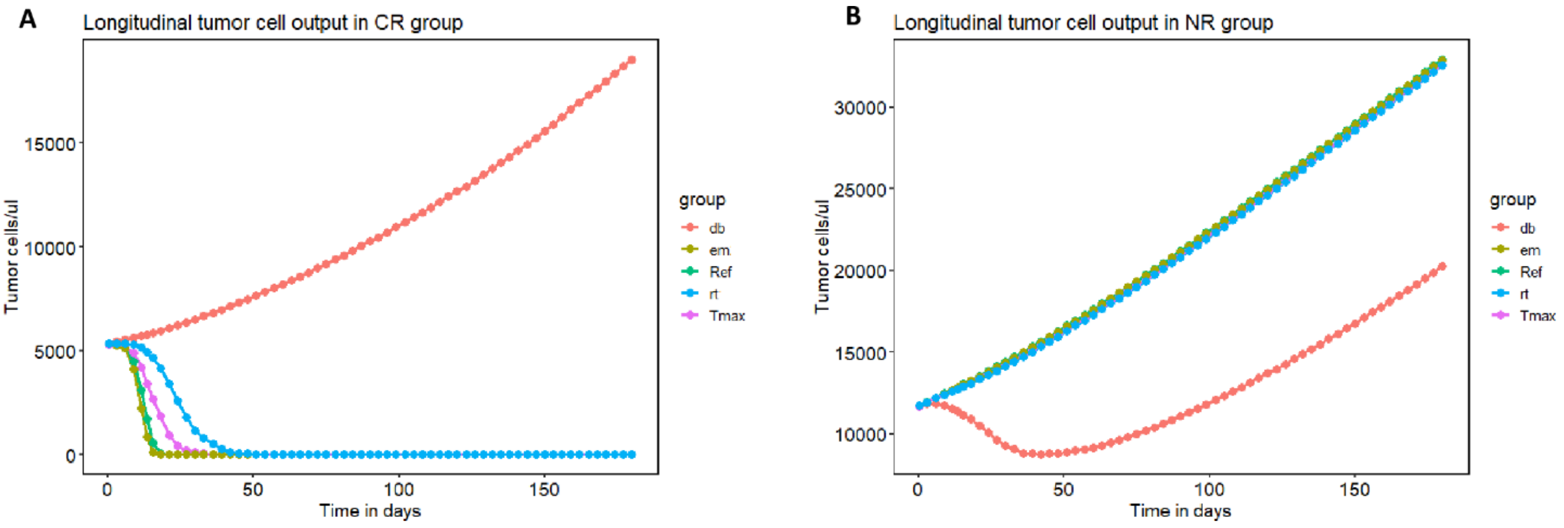


**Supplementary figure 3.** Median longitudinal tumor cell predictions from simulating 1000 CR and NR virtual patients each, over the period of 180 days after varying one parameter at a time. In comparison with the reference curve (green) where the estimated population parameters for CR or NR from model fitting were used respectively, $d_b$ (killing rate of B-cells by CART cells) parameter shows greatest impact on tumor cell dynamics.

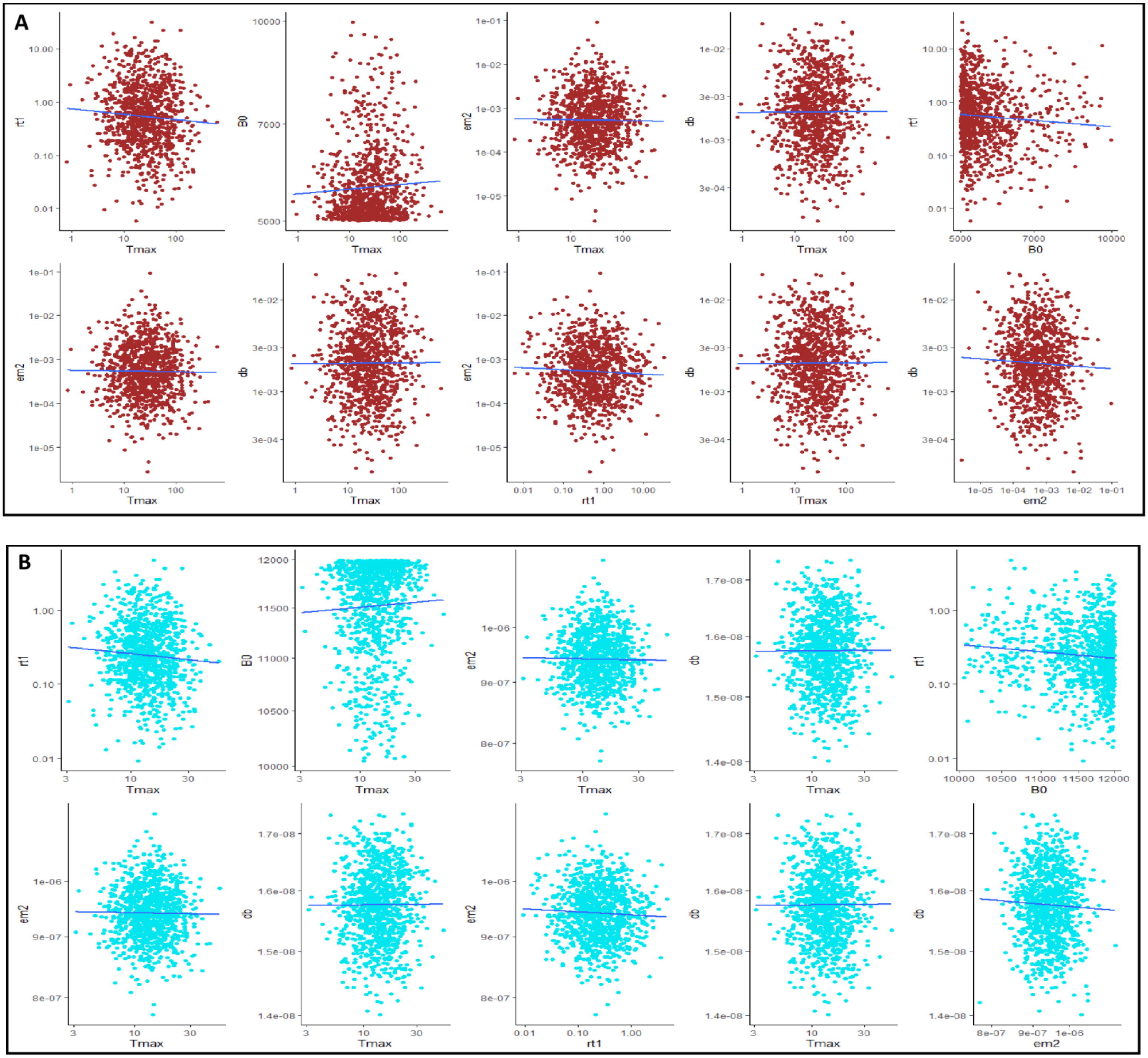


**Supplementary figure 4.** Correlations between parameters in (A) CR patients (red) and (B) NR Patients (blue). Except for a small negative correlation between CAR T cell proliferation rate ($r_t$) and initial tumor burden ($B_0$) in NR group, none of these correlations were statistically significant.

### Monolix Model Code:

```
[LONGITUDINAL]

input = {F,Tmax,IL60,B0,rt1,de2,em2,db,pi2,di}

IL60 = {use = regressor}

PK:

V = 3e6

depot(target=E, p=F/V)

EQUATION:

odeType=stiff

t_0 = 0

E_0 = 0

I_0  = IL60

B_0 = B0

pi1 = IL60 * di

rb = 9.3e-3

Kb = 57000

dm = 0.00027

me2 = 1

de1=0

rt2=0

em1=0

me1=0

if t<= Tmax

rt=rt1

de = de1

em = em1

me = me1
```

```
else

rt=rt2

de =de2

em = em2

me = me2

end

Bf = B/(hb+B)

ddt_B  =   rb * B*(1-B/Kb) - db * B * E

ddt_E =  rt*Bf* E - de*E - (1-Bf)*em*E + Bf*me*M

ddt_M = (1-Bf)*em*E - dm*M - Bf*me*M

ddt_I   = -di*I + pi2 *  B * E + pi1

T =  E+M

Tumor = B

OUTPUT:

output = {T,Tumor,I}

table = {T,Tumor,E,M,I}
```
